# Longitudinal associations of psychological distress with subsequent cognitive decline and dementia: a multi-cohort study

**DOI:** 10.1101/2025.08.19.25333919

**Authors:** J. Stafford, S. Dekhtyar, T. C Russ, A. Singh-Manoux, J. Maddock, K. Walters, V. Orgeta, N. Davies, J. B. Kirkbride, M. Richards, R. Howard, P. Patalay

## Abstract

**Background:** Psychological distress has been linked with later cognitive impairment and dementia, although the nature of the association remains unclear. Using a multi-cohort approach, we examined longitudinal associations of psychological distress with subsequent cognition and dementia, testing whether findings varied by age of assessment, severity, and persistence of psychological distress.

**Methods:** We used five longitudinal studies: Caerphilly Prospective Study, English Longitudinal Study of Ageing, National Child Development Study, National Survey of Health and Development, and Whitehall II. We examined associations with changes in cognition using linear and mixed effects models, and dementia using logistic regression. Results were pooled using two-stage individual participant data meta-analysis.

**Findings:** Pooled analyses (total N=24,564) showed that greater psychological distress was associated with lower subsequent cognitive level (β=−0.03 [95% CI: −0.06; −0.01]; I2=69.7%). Associations were present for clinically-significant (β=−0.06 [−0.12; −0.00]; I2=62.2%), persistent (β=−0.12 [−0.23; −0.02]; I2=82.5%) and intermittent distress (β=−0.09 [−0.12; −0.05]; I2=0%). Baseline distress was not associated with rates of subsequent cognitive decline. Psychological distress was associated with subsequent dementia (OR=1.12 [1.04; 1.20]; I2=0%), including for clinically-significant (OR=1.28 [1.07; 1.52; I2=0%]), persistent (OR=1.43 [1.04; 1.98]; I2=44.0%) and intermittent symptoms (OR=1.32 [1.02; 1.71]; I2=40.3%). Dementia was associated with psychological distress assessed in later life (age 65-75 OR: 1.29 [1.18; 1.40]; I2=13.9%), but not mid-life (age 45-54 OR: 1.09 [0.93; 1.28]; I2=34.0%).

**Interpretation:** In this multi-cohort study, psychological distress was associated with subsequent dementia and lower subsequent cognitive level, including for both persistent and intermittent distress. Associations with dementia were present when distress was assessed at older ages but not at ages 45-54 years, suggesting that associations might partly represent early preclinical markers of dementia neuropathology. Findings highlight the potential relevance of psychological distress in informing dementia prevention and supporting identification of high-risk groups, both of which are major global public health priorities.

## Background

Experiencing mental health difficulties, such as depression, in mid- and late-life has been associated with subsequent cognitive impairment and dementia.^1,2^ However, few studies have examined cognitive outcomes in broader psychological distress, which refers to symptoms of depression, anxiety, stress and somatic complaints that are not severe enough to meet diagnostic criteria for a psychiatric disorder.^3^ In addition, temporality and the mechanisms underlying these associations are poorly understood. Gaining further insight into longitudinal relationships between psychological distress, cognition and dementia could help to inform dementia prevention approaches, and earlier identification of high-risk groups, both of which are major priority areas for global public health.

Most research to date has focused on cognitive outcomes in relation to specific in psychiatric diagnoses and symptoms, particularly depression,^1^ which was identified as a modifiable risk factor in the Lancet Dementia commission.^4^ While broader psychological distress has also been linked with dementia,^5–8^ the evidence is relatively sparse, and has been limited by varying measures of distress, including the use of single questions.^9,10^

Aspects of psychological distress have also shown associations with poorer subsequent cognitive functioning^11,12^ and decline,^2^ although findings regarding cognitive trajectories have been mixed.^13,14^ For instance, in the 1946 birth cohort, emotional symptoms in adolescence, but not adulthood, were associated with poorer subsequent cognitive functioning at age 43, but not with decline between ages 43-69 years.^15^

In addition, the temporal nature of these associations remains poorly understood. In particular, it is unclear whether psychological distress symptoms are causal risk factors for dementia, or early markers of dementia neuropathology during the preclinical period (reverse causality). Many previous studies have been limited due to short follow-up periods (<10 years), where any association could reflect reverse causality. Studies with longer follow-up periods have yielded mixed results, with several finding longstanding links between mid-life psychological distress and dementia.^7^ In contrast, other studies have only found associations with late-life depression, and closer to dementia diagnosis (~11 years before).^16,17^

Further research is also needed to examine the role of severity and persistence of psychological distress symptoms, where findings been mixed. For instance, in depression, some studies have identified associations for mild and subsyndromal symptoms,^18,19^ while others only found associations with more severe symptoms.^20,21^ There is some evidence that associations may vary by sex.^17^ For instance, a Norwegian population-based study found longstanding associations between affective symptoms and dementia in women, whereas for men, associations were only found closer to dementia diagnosis.^17^ However, evidence on sex differences in limited, and few studies have examined variation in relation to other sociodemographic factors, such as education level.

To address these gaps, using multi-cohort longitudinal data, we investigated longitudinal relationships between psychological distress and subsequent cognitive trajectories and dementia risk. We sought to gain further insight into the temporal nature of these relationships by using studies with long follow-up periods and measures of psychological distress from both early-to mid-adulthood and mid-to late-adulthood. We examined whether findings varied by: age of psychological distress assessment, severity and persistence of psychological distress, and sociodemographic factors including sex, education level and occupational social class.

## Methods

### Pre-registration

We pre-registered our protocol on the Open Science Framework (OSF) platform (https://doi.org/10.17605/OSF.IO/ZHG8B). Amendments to the protocol are reported in Table S1.

#### Study design and participants

We examined associations between psychological distress, cognition and dementia using a multi-cohort approach, and included data from five longitudinal studies (Table S2): the Caerphilly Prospective Study (CAPS), the English Longitudinal Study of Ageing (ELSA), Whitehall II (WHII), the National Child Development Study (NCDS); and the National Survey of Health and Development (NSHD). All data sources were accessed through the Dementias Platform UK (DPUK). Subsequent fluid cognition outcomes were assessed at a single time point in all datasets, and in NSHD, WHII and ELSA, we examined associations with cognitive trajectories. We examined dementia outcomes in all studies, except NCDS. Within each study, our analytical sample included participants with data on psychological distress and without prevalent dementia at baseline (where information on dementia was available at baseline); with assessments of cognition in the single time point used in analyses; and at least two cognitive assessments after baseline psychological distress assessment where cognitive trajectories were examined.

A lived experience advisory group of three former carers for people with dementia were involved throughout the study. The group provided input on study conceptualisation, protocol development and interpretation of findings, and will also be involved in shaping plans for dissemination.

#### Psychological distress exposures

Self-reported psychological distress at baseline was measured using validated assessment tools within each study, as reported in Table S3 and Supplementary Information. In WHII, psychological distress was first measured in 1985, when participants were aged 35-55 years, using the General Health Questionnaire (GHQ-30),^22^ which is widely used to screen for psychiatric disorders in community samples. In CAPS, psychological distress was assessed using the GHQ-30 assessment in wave 1, in which participants were aged 45-59 years. In ELSA, psychological distress at wave 1 was assessed using the 8-item Centre for Epidemiological Studies Depression scale (CES-D),^23^ which measures symptoms including depressed mood, restless sleep, and decreased energy and enjoyment in life. In NCDS, psychological distress at age 23 was assessed using the Malaise Inventory,^24^ a measure of emotional disturbance and associated somatic symptoms.

In NSHD, psychological distress was assessed in adulthood at the following ages: age 36 (Present State Examination),^25^ age 43 (Psychiatric Symptom Frequency),^26^ and at ages 53, 60-64 and 68-70 years (GHQ-28).^27^ To allow comparison across waves, in line with guidance from CLS and CLOSER,^28^ we created harmonised variables based on seven conceptually similar items capturing psychological distress consistently within each measure, with items pertaining to low mood, fatigue, tense/stressed, sleep problems, panic, hopelessness and health anxiety (Supplementary Information).

In primary analyses, baseline psychological distress in adulthood was used as the main exposure. We also examined associations with clinically-significant psychological distress using binary variables based on validated cut-off scores on questionnaires within each study (Table S3; Supplementary Information). In addition, we examined whether associations varied depending on whether clinically-significant psychological distress was observed persistently prior to cognitive assessments during the first three waves in which psychological distress assessments were available in adulthood (high symptoms in two or three waves) versus intermittently high (high symptoms in one wave only), relative to those without clinically-significant psychological distress.

#### Outcomes

We computed an overall level of fluid cognition based on individual cognitive assessments of domains including memory, verbal fluency and speed of processing. To facilitate comparison across studies and tests, we standardised each cognitive test across timepoints and within studies on a common standard deviation-based scale, with a mean of 0 and a standard deviation of 1. General fluid cognitive level was computed by averaging standardised scores across individual domains and re-standardising the fluid cognitive level. Full information about assessments of individual cognitive domains within each study is provided in Supplementary Information.

We also examined dementia in CAPS, ELSA, WHII and NSHD. In WHII, dementia diagnoses were obtained from NHS Digital’s Hospital Episode Statistics based on ICD-10 codes F00, F01, F03, G30 and G31, with 78% sensitivity and 92% specificity,^29^ and from self-reported dementia diagnosis. In ELSA, dementia was ascertained from self-reported physician diagnosis, with additional cases obtained from the Informant Questionnaire on Cognitive Decline in the Elderly (IQCODE), which has been validated as a sensitive screening tool for dementia,^30^ based on a validated cut-off score of 3.5 or above.^31^ In CAPS, at wave V (aged 68-82 years) the Cambridge Cognitive Examination (CAMCOG),^32^ a standardized instrument to measure dementia and assess cognitive impairment, was used to select people for detailed clinical assessment, from which participants were classified as having normal cognition, cognitive impairment without dementia, or dementia. Because of the smaller sample in wave V, we included those with cognitive impairment or dementia in our outcome. In NSHD, at age 77 years the AD8 interview was administered to participants’ informants to distinguish those with very mild dementia from those without dementia.^33^ The AD8 has demonstrated strong correlation with Clinical Dementia Rating Domains and performance on neuropsychological tests.^34^ Further detail about dementia ascertainment is provided in Supplementary Information.

#### Covariates

Where available across studies, models were adjusted for the following covariates at baseline: age (continuous in years), sex (male, female), education level (GCSE/O-level or equivalent, less than GCSE/O-level), occupational position (manual, nonmanual, other/none), marital and cohabitation status (married and/or living with a partner, unmarried and living alone), long-term health conditions (none, one or more), smoking (current smoker, ex-smoker, never smoked), alcohol consumption (low, moderate, high), physical activity (low, moderate, high), baseline or childhood cognition where possible (for cognitive outcomes). Full information about covariates within each study is provided in Supplementary Information.

#### Statistical analyses

We used linear regression models to examine associations between baseline psychological distress and subsequent standardised fluid cognitive level assessed at a single time point (mean ages 62-72 years). In addition, in ELSA, WHII and NSHD, where assessments of cognition were available in three or more time points, we used linear mixed models with random slope and intercept to examine associations between baseline psychological distress and subsequent trajectories of standardised fluid cognitive level over time, with time centred on baseline date and coded in years.

We investigated associations between baseline psychological distress and subsequent dementia in CAPS, ELSA, WHII, and NSHD using logistic regression models. We examined whether associations with dementia varied by age of psychological distress assessment, defining three age bands for mental health exposure: ages 45-54; 55-64; and 65-75 years. Within each study, we selected the psychological distress score from the assessment closest to the target age of 50, 60, or 70 years, respectively.

For all analyses, we presented findings from models partially adjusted for age and sex and maximally adjusted for as many available covariates from the list above. We used interaction terms between psychological distress and covariates to examine whether associations varied by sociodemographic factors, including: sex in all datasets, except CAPS, where data are only available for men, education level (GCSE/O-level or equivalent vs less than GCSE/O-level), and occupational social class (manual, non-manual, other/none). In secondary analyses, we examined associations with cognitive outcomes in relation to depressive and anxiety symptoms individually. We examined associations between baseline psychological distress and subsequent dementia using Cox regression as a sensitivity analysis in ELSA and WHII, where date of dementia diagnosis was available.

We analysed data using an individual participant data two-stage meta-analysis approach, with study-specific analyses completed in the first step, followed by pooling study-specific estimates across studies where appropriate. Before analysis, we harmonised all variables between studies as far as possible (Supplementary Information). We assessed heterogeneity between estimates using the I2 statistic. We applied multiple imputation models using chained equations to impute missing covariate data (Table S4). All analyses were completed in Stata version 18.0

## Results

Baseline characteristics of participants within each study in our analyses are presented in Table 1. A participant flow diagram is provided in Figure S1, indicating participants excluded due to having dementia at baseline, missing information on baseline psychological distress, or without information on either cognition in the single time point used in analyses or at least two cognitive assessments after baseline psychological distress assessment. Participants included in the analysis differed on socioeconomic and health-related factors from those excluded. For instance, those excluded were more likely to have lower education, manual occupations, comorbidities, and to have negative health behaviours (Table S5). The overall analytical sample across studies was 24,564 (44.4% women; pooled mean age at baseline: 43.8 years).

**Table 1.**
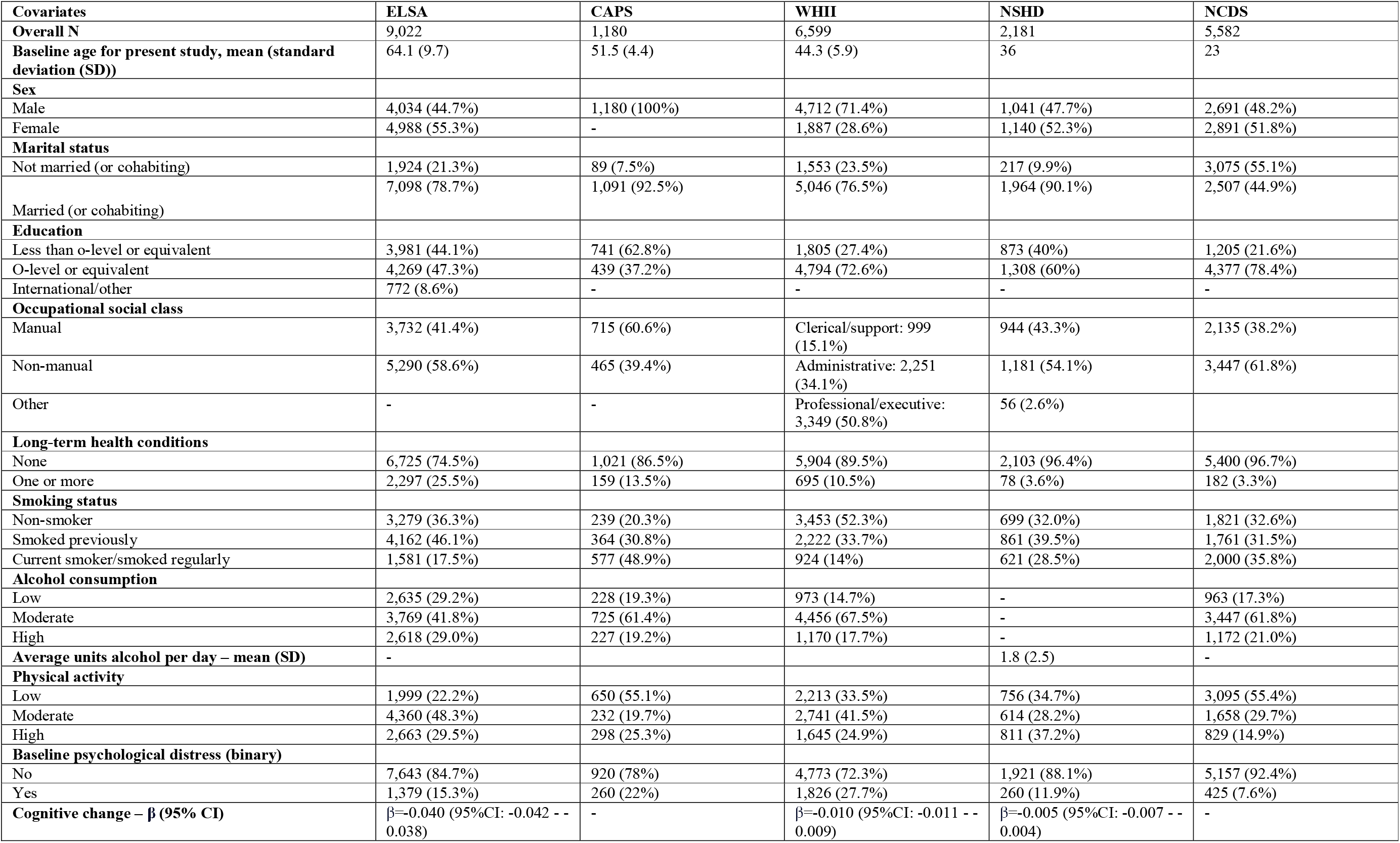
Descriptive statistics for psychological distress exposures, cognitive outcomes and covariates.

| Covariates | ELSA | CAPS | WHII | NSHD | NCDS |
| --- | --- | --- | --- | --- | --- |
| **Overall N** | 9,022 | 1,180 | 6,599 | 2,181 | 5,582 |
| **Baseline age for present study, mean (standard deviation (SD))** | 64.1 (9.7) | 51.5 (4.4) | 44.3 (5.9) | 36 | 23 |
| **Sex** |  |  |  |  |  |
| Male | 4,034 (44.7%) | 1,180 (100%) | 4,712 (71.4%) | 1,041 (47.7%) | 2,691 (48.2%) |
| Female | 4,988 (55.3%) | - | 1,887 (28.6%) | 1,140 (52.3%) | 2,891 (51.8%) |
| **Marital status** |  |  |  |  |  |
| Not married (or cohabiting) | 1,924 (21.3%) | 89 (7.5%) | 1,553 (23.5%) | 217 (9.9%) | 3,075 (55.1%) |
| Married (or cohabiting) | 7,098 (78.7%) | 1,091 (92.5%) | 5,046 (76.5%) | 1,964 (90.1%) | 2,507 (44.9%) |
| **Education** |  |  |  |  |  |
| Less than o-level or equivalent | 3,981 (44.1%) | 741 (62.8%) | 1,805 (27.4%) | 873 (40%) | 1,205 (21.6%) |
| O-level or equivalent | 4,269 (47.3%) | 439 (37.2%) | 4,794 (72.6%) | 1,308 (60%) | 4,377 (78.4%) |
| International/other | 772 (8.6%) | - | - | - | - |
| **Occupational social class** |  |  |  |  |  |
| Manual | 3,732 (41.4%) | 715 (60.6%) | Clerical/support: 999 (15.1%) | 944 (43.3%) | 2,135 (38.2%) |
| Non-manual | 5,290 (58.6%) | 465 (39.4%) | Administrative: 2,251 (34.1%) | 1,181 (54.1%) | 3,447 (61.8%) |
| Other | - | - | Professional/executive: 3,349 (50.8%) | 56 (2.6%) |  |
| **Long-term health conditions** |  |  |  |  |  |
| None | 6,725 (74.5%) | 1,021 (86.5%) | 5,904 (89.5%) | 2,103 (96.4%) | 5,400 (96.7%) |
| One or more | 2,297 (25.5%) | 159 (13.5%) | 695 (10.5%) | 78 (3.6%) | 182 (3.3%) |
| **Smoking status** |  |  |  |  |  |
| Non-smoker | 3,279 (36.3%) | 239 (20.3%) | 3,453 (52.3%) | 699 (32.0%) | 1,821 (32.6%) |
| Smoked previously | 4,162 (46.1%) | 364 (30.8%) | 2,222 (33.7%) | 861 (39.5%) | 1,761 (31.5%) |
| Current smoker/smoked regularly | 1,581 (17.5%) | 577 (48.9%) | 924 (14%) | 621 (28.5%) | 2,000 (35.8%) |
| **Alcohol consumption** |  |  |  |  |  |
| Low | 2,635 (29.2%) | 228 (19.3%) | 973 (14.7%) | - | 963 (17.3%) |
| Moderate | 3,769 (41.8%) | 725 (61.4%) | 4,456 (67.5%) | - | 3,447 (61.8%) |
| High | 2,618 (29.0%) | 227 (19.2%) | 1,170 (17.7%) | - | 1,172 (21.0%) |
| **Average units alcohol per day – mean (SD)** | - |  |  | 1.8 (2.5) | - |
| **Physical activity** |  |  |  |  |  |
| Low | 1,999 (22.2%) | 650 (55.1%) | 2,213 (33.5%) | 756 (34.7%) | 3,095 (55.4%) |
| Moderate | 4,360 (48.3%) | 232 (19.7%) | 2,741 (41.5%) | 614 (28.2%) | 1,658 (29.7%) |
| High | 2,663 (29.5%) | 298 (25.3%) | 1,645 (24.9%) | 811 (37.2%) | 829 (14.9%) |
| **Baseline psychological distress (binary)** |  |  |  |  |  |
| No | 7,643 (84.7%) | 920 (78%) | 4,773 (72.3%) | 1,921 (88.1%) | 5,157 (92.4%) |
| Yes | 1,379 (15.3%) | 260 (22%) | 1,826 (27.7%) | 260 (11.9%) | 425 (7.6%) |
| **Cognitive change – $\beta$ (95% CI)** | $\beta = -0.040$ (95% CI: -0.042 - -0.038) | - | $\beta = -0.010$ (95% CI: -0.011 - -0.009) | $\beta = -0.005$ (95% CI: -0.007 - -0.004) | - |

### Psychological distress and general fluid cognition

First, we investigated associations between baseline psychological distress and general fluid cognitive level at a single time point (mean ages 62-72 years; Table 2; Figure 1; Figures S2-S3). Pooled analyses across five studies showed that greater psychological distress was associated with poorer subsequent standardised fluid cognitive level in fully adjusted models (β=−0.03 [95% CI: −0.06; −0.01]; I2=69.7%). This pattern was also observed using cut-off scores indicating clinically-significant psychological distress (fully adjusted model (β=−0.06 [−0.12; −0.00]; I2=62.2%). Associations were found for persistent (β=−0.12 [−0.23; −0.02]; I2=82.5%) and intermittent distress (β=−0.09 [−0.12; − 0.05]; I2=0%).

**Table 2.**
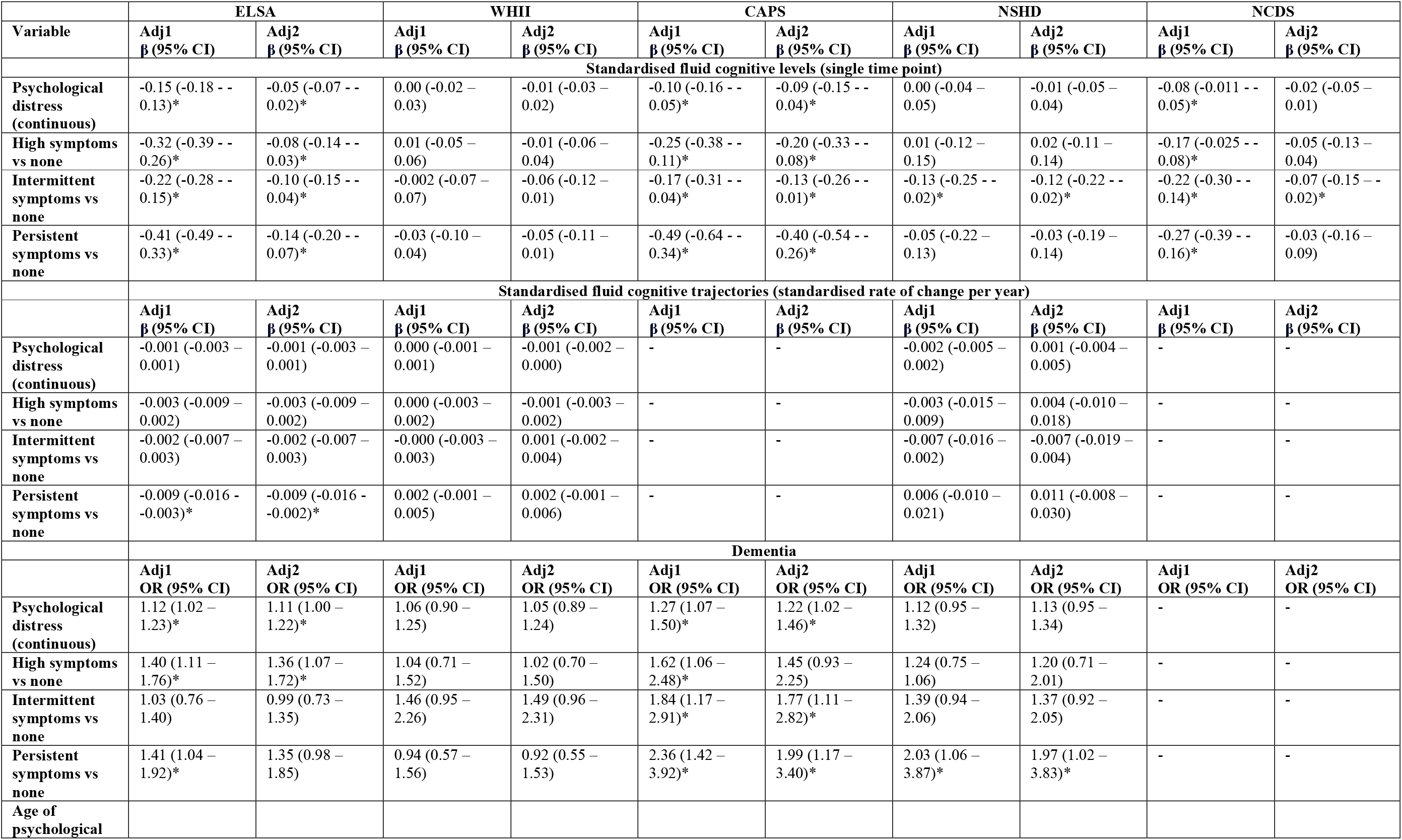
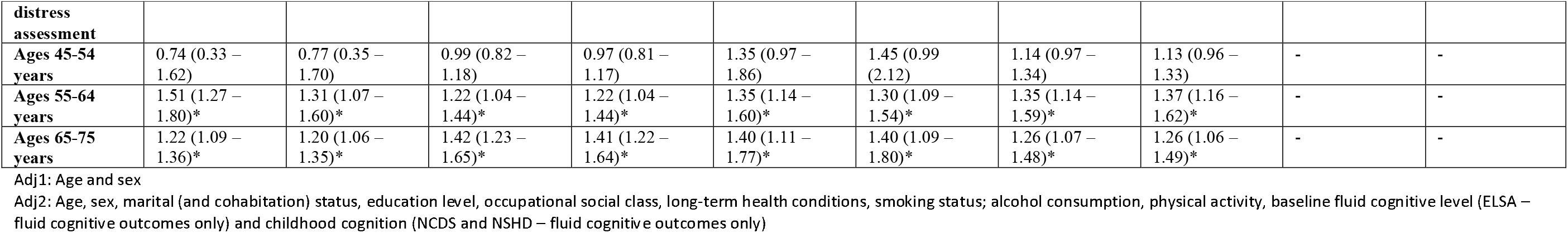
Associations between psychological distress and cognitive outcomes.

**Figure 1.**
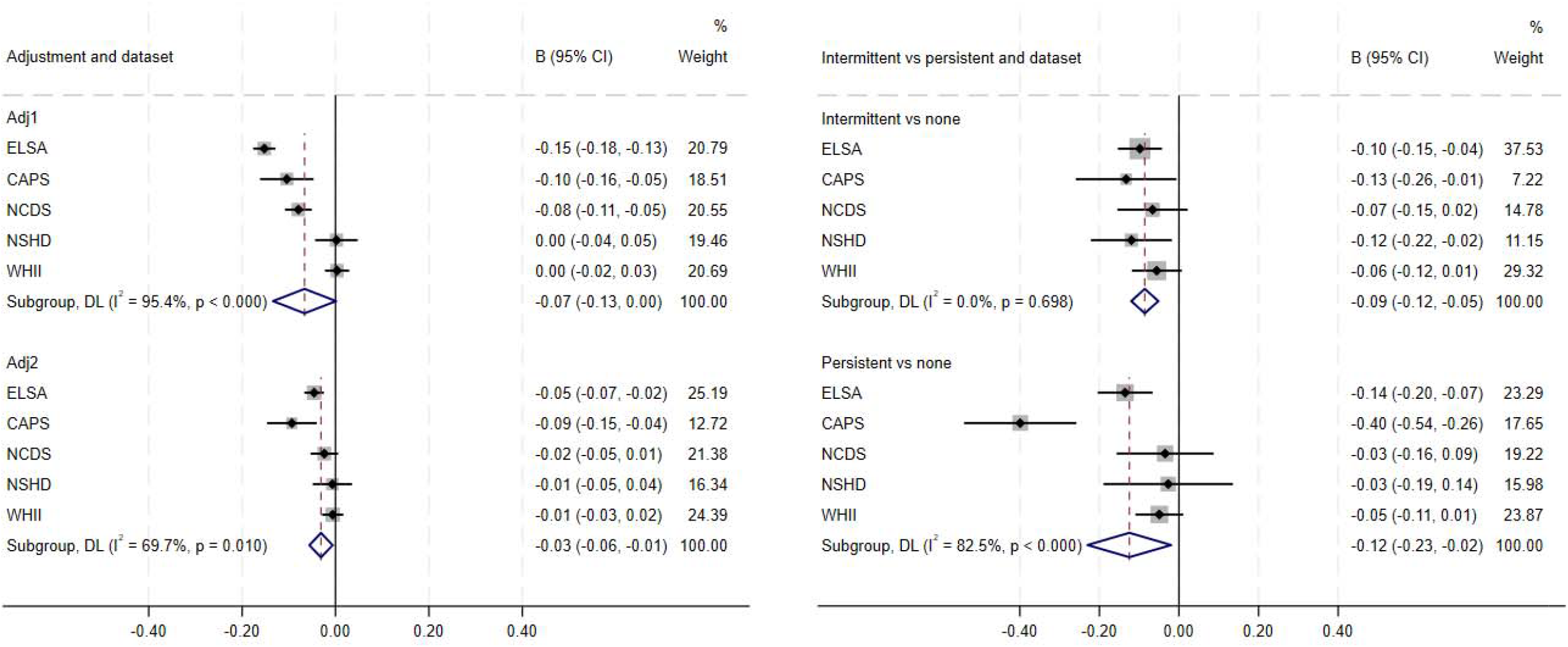
Pooled association between psychological distress and subsequent fluid cognitive levels – overall association with base distress (left); intermittently and persistently high vs no high psychological distress (right)

Second, we examined associations between psychological distress and subsequent general fluid cognitive change in ELSA, NSHD and WHII (Table 2; Figure S4). We observed decline in cognition over time in all three studies (ELSA: β=−0.040 [−0.042; −0.038]); NSHD (β=−0.005 [−0.007 − −0.004]); WHII (β=−0.010 [−0.011 − −0.009]; Table 1). When data were pooled across studies, baseline psychological distress, including clinically-significant distress, was not associated with subsequent cognitive trajectories. In ELSA, persistent (β=−0.009 [−0.016; −0.002]), but not intermittent distress was associated with cognitive decline, but this was not observed in other studies.

### Psychological distress and dementia

Third, we examined associations between psychological distress and subsequent dementia in CAPS, ELSA, WHII and NSHD (Table 2; Figures 2–3; Figure S5-S7). Pooling estimates across studies, we found evidence of an association between baseline psychological distress and subsequent dementia (fully adjusted OR=1.12 [1.04; 1.20]; I2=0%). This was also observed for clinically-significant symptoms (fully adjusted OR=1.28 [1.07; 1.52; I2=0%]). Both persistent (OR=1.43 [1.04; 1.98]; I2=44.0%) and intermittent distress were associated with subsequent dementia (OR=1.32 [1.02; 1.71]; I2=40.3%). We also examined variation in associations across the following age bands, where estimates could be pooled across studies: ages 45-54; 55-64; and 65-75 years. Pooled associations between psychological distress and dementia were found when psychological distress was assessed at ages 65-75 years (OR: 1.29 [1.18; 1.40]; I2=13.9%) and 55-64 years (OR: 1.29 [1.18; 1.41]; I2=0%), but not at ages 45-54 years (OR: 1.09 [0.93; 1.28]; I2=34.0%).

**Figure 2.**
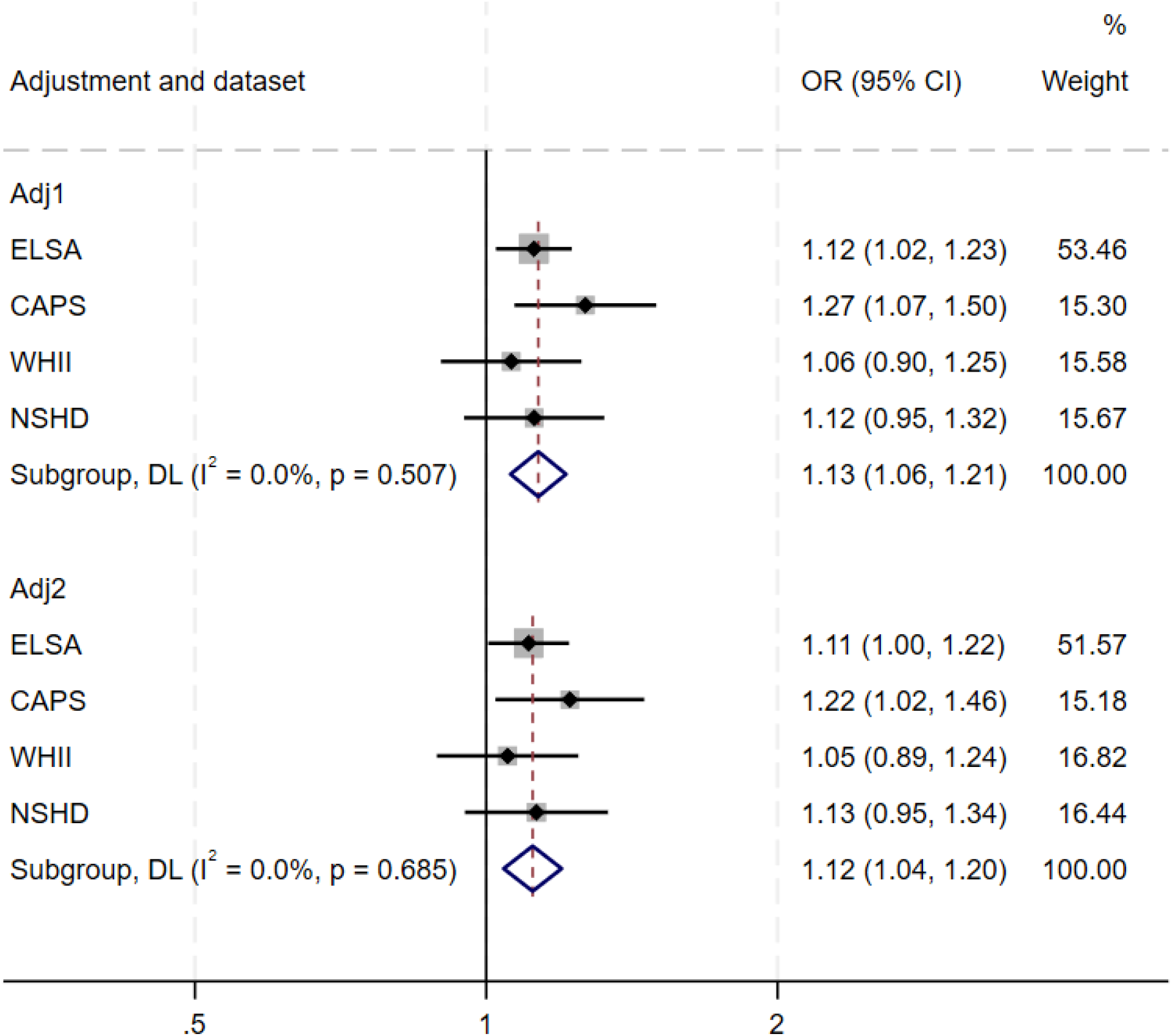
Pooled association between baseline continuous psychological distress and subsequent dementia

**Figure 3.**
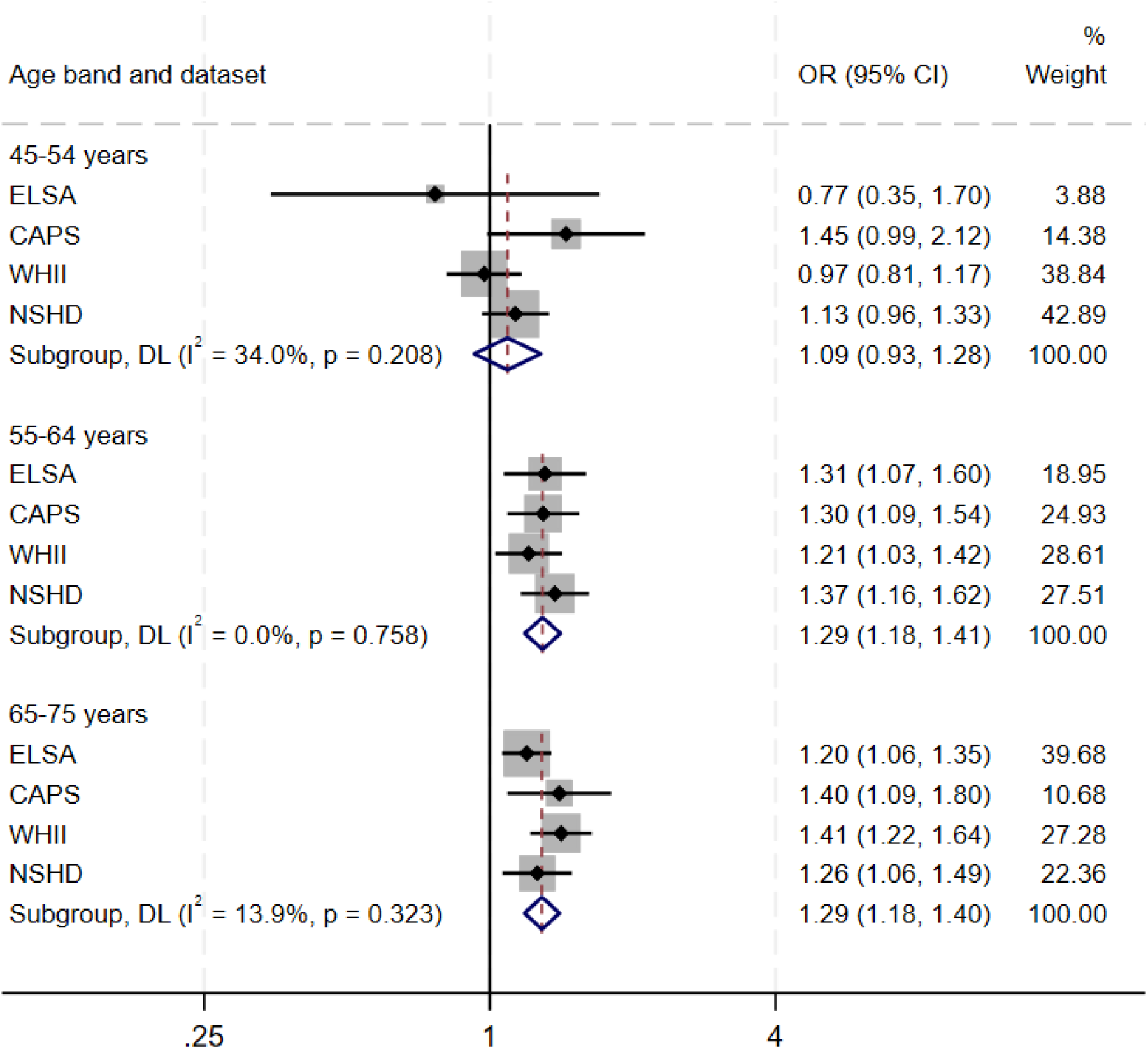
Pooled association between psychological distress and dementia by age of psychological distress assessment

### Sensitivity and secondary analysis and subgroup differences

Patterns of association were similar when examining depression and anxiety symptoms individually (Figure S3, Figure S7). We did not find evidence of effect modification by sex, education level, or occupational social class. Similar patterns of association were found using Cox regression models to examine dementia outcomes in ELSA and WHII, where information on timing of dementia diagnosis was available. In ELSA, baseline psychological distress was associated with subsequent dementia (fully adjusted hazard ratio (HR): 1.16 [1.06; 1.27]), and consistent with logistic regression models, baseline distress was not associated with dementia in WHII (HR: 1.02 [0.86; 1.20]).

## Discussion

### Summary and meaning of findings

In this multi-cohort study, psychological distress was found to be associated with subsequent dementia and lower fluid cognitive level, but not with decline in cognition over time. Associations were found across the full range of psychological distress symptoms, including clinically-significant symptoms. Both intermittent and persistent symptoms were associated with poorer cognitive outcomes. Associations with dementia were present for psychological distress assessed at ages 55-64 and 65-75 years, but not at ages 45-54 years.

Findings indicate that not only severe psychological distress, but also broader and subclinical aspects of distress are associated with poorer subsequent cognitive outcomes. In line with this, previous studies have shown associations between mild and subsyndromal depressive symptoms and subsequent dementia.^18,19^ In contrast, other studies have only identified associations with dementia for severe depression symptoms.^20,21^ In line with previous studies, we found that persistent psychological distress was associated with poor cognitive outcomes,^35,36^ although we also found associations for intermittent distress. These findings could have important implications for cognitive health at a population level, given the high prevalence of mild, intermittent psychological distress symptoms across the life course.^37^

In secondary analyses, both depression and anxiety symptoms showed associations with poorer cognitive outcomes. Similar patterns of association found may reflect the substantial overlap and high comorbidity between depressive and anxiety symptoms.^38^ Despite this, previous findings relating to anxiety have been mixed, with several studies showing longitudinal links between anxiety and subsequent dementia,^17,39–42^ while others did not observe associations with cognitive outcomes.^43,44^ These findings could help to inform approaches to dementia prevention, including the updated Lancet dementia commission, given that anxiety was noted as a potential modifiable risk factor, but was not included in the final model due to insufficient supporting evidence.^4^

We did not find evidence of variation by sex, education level, or occupational class. This contrasts a previous Norwegian cohort study which reported differences by sex, whereby no difference in dementia risk was found among those with anxiety and depression 11 years before dementia overall, while in women, a difference was present 33 years before dementia diagnosis.^17^

Mechanisms linking psychological distress with subsequent cognitive outcomes remain unclear. Distress could cause poorer subsequent cognitive outcomes, either directly or via pathways such as reduced cognitive and brain reserve,^45^ and those involving chronic stress.^46^ Psychological distress has also shown links with systemic inflammation,^47^ which has been implicated in the pathophysiology of dementia.^48^ Psychological distress is also associated with poor cardiometabolic health^49^ and negative health behaviours,^50^ which have been identified as major risk factors for dementia.^4^ On the other hand, shared genetics and environmental and health-related factors could shape both psychiatric and cognitive outcomes.^16^

Associations between psychological distress, cognition and dementia could also reflect reverse directionality, given that neuropathology in the preclinical period of dementia unfolds progressively over several decades before cognitive and functional symptoms are apparent.^51,52^ Hence, psychiatric symptoms observed closer to dementia diagnosis could reflect early markers of neuropathology, rather than causal risk factors.

In the present study, associations were observed when psychological distress was assessed between ages 55-64 and 65-75 years, but not at age 45-54 years. Our findings align with several previous studies reporting stronger associations with mental health difficulties in later-life and over shorter follow-up periods.^53–55^ Some studies only identified associations over shorter follow-up periods (~11 years before dementia), with associations not observed over longer durations.^16,17^ Taken together, findings suggest that the association between psychological distress and subsequent dementia may partly reflect reverse directionality.

These findings have implications for informing preventative models for dementia, and for clinical practice. In particular, if psychological distress symptoms represent early signs of underlying neuropathology, this information could play a role in identifying people who may be at higher risk of adverse cognitive outcomes at an earlier stage, indicating a need for increased monitoring for cognitive and functional decline, and guiding referral pathways.

However, it is important to note that several previous studies have observed smaller, longstanding associations between depression and dementia across multiple decades,^6,7,17^ suggesting that findings are unlikely to be fully explained by reverse causality. Further, in a Norwegian population-based cohort, stronger associations with dementia were found for psychological distress assessed in early-mid-life compared to later mid-life.^7^

### Strengths and limitations

Strengths of the study include the multi-cohort approach involving five UK longitudinal studies, allowing a large overall sample size. Included studies had long follow-up periods, with repeated measures of psychological distress and cognitive outcomes over many years, and rich information on a range of relevant sociodemographic and health-related covariates.

There are several limitations to note. While we sought to harmonise exposures, outcomes and covariates as far as possible between studies, it remains possible that heterogeneity in estimates between studies could reflect differences in measurement across studies. Variation in findings between studies could also reflect different study settings, length of follow-up time, and ages at selected baseline assessments, where experiences and effects of psychological distress on later cognitive outcomes may differ. Assessments of dementia varied between studies, with our outcome in CAPS also including people with cognitive impairment without dementia, and the AD8 in NSHD being used to identify those with very mild dementia symptoms. CAPS and NSHD also lacked information on timing of dementia diagnosis, limiting inference about temporal relationships with psychological distress. It should be noted that CAPS only included men. Not all studies had measures of dementia or repeated measures of cognitive outcomes, hence it was only possible to examine dementia and cognitive decline outcomes in a smaller subset of studies.

## Conclusion

Applying a multi-cohort approach, we found that psychological distress was associated with poorer subsequent fluid cognitive level and dementia, but not with cognitive trajectories over time. These associations were not restricted to persistent, clinically-significant distress, but were found across the full range of psychological distress symptoms, including for intermittent symptoms. Our findings of stronger associations with distress assessed at older ages, taken alongside the wider literature on temporal relationships, indicate that at least some of the association between psychological distress and dementia may be explained by reverse causality. However, smaller longstanding associations between psychiatric symptoms and dementia in the literature indicate that reverse causality is unlikely to fully explain these relationships.

Further research is needed to tease apart whether longstanding relationships reflect causal effects, or shared genetic and environmental causes. In addition, there is a need to examine dementia risk in relation to a wider range of psychiatric symptoms, and to explore potential underlying mechanisms. Findings could have important implications for informing dementia prevention approaches. In addition, for psychological distress observed closer to dementia, which may reflect reverse causality, this information could contribute to identifying individuals at higher risk for poor cognitive outcomes at an earlier stage, potentially helping to guide referral pathways and inform clinical management.

## Data Availability

DPUK provided access to all datasets included in this study through MRC grant ref MR/L023784/2 (core funding).
ELSA data used in this study are available to download through the UK Data Service.
Data for NCDS (SN 6137) are available through the UK Data Service.
NSHD data are available on request to the NSHD Data Sharing Committee. Interested researchers can apply to access the NSHD data via a standard application procedure.
Caerphilly Prospective Study data can be accessed following review and approval by the steering committee.
Researchers interested in accessing the Whitehall II data can apply through the Dementias Platform UK or the Whitehall Scientific Committee.

## Funding acknowledgements

JS was supported by an Alzheimer’s Society Postdoctoral Fellowship. The English Longitudinal Study of Ageing was developed by a team of researchers based at University College London, NatCen Social Research, the Institute for Fiscal Studies, the University of Manchester, and the University of East Anglia. The data were collected by NatCen Social Research. The funding is currently provided by the National Institute on Aging (Ref: R01AG017644) and by a consortium of UK government departments: Department for Health and Social Care; Department for Transport; Department for Work and Pensions, which is coordinated by the National Institute for Health Research (NIHR, Ref: 198-1074). Funding has also been provided by the Economic and Social Research Council (ESRC). 1958 National Child Development Study are supported by the Centre for Longitudinal Studies, Resource Centre 2015-20 grant (ES/M001660/1) and a host of other co-funders. The 1946 NSHD cohort is hosted by the MRC Unit for Lifelong Health and Ageing funded by the Medical Research Council (MC_UU_00019/1Theme 1: Cohorts and Data Collection). The Whitehall II study has been supported by grants from the British Medical Research Council (MR/K013351 and MR/ R024227/1); the British Heart Foundation (PG/11/63/29011 and RG/13/2/30098); the British Health and Safety Executive; the British Department of Health; the National Heart, Lung, and Blood Institute (R01HL036310); the National Institute on Aging, National Institute of Health (R01AG013196, R01 AG034454); the Economic and Social Research Council (ES/ J023299). CaPS was funded by the Medical Research Council (grant G9824960-E01/1), and phase 5 was funded by a grant from the Alzheimer’s Society.

## Acknowledgements

We acknowledge participants and the research teams involved in NSHD, NCDS, ELSA, WHII and CAPS for their contribution to this research.

## Data sharing

DPUK provided access to all datasets included in this study through MRC grant ref MR/L023784/2 (core funding). ELSA data used in this study are available to download through the UK Data Service. Data for NCDS (SN 6137) are available through the UK Data Service. NSHD data are available on request to the NSHD Data Sharing Committee. Interested researchers can apply to access the NSHD data via a standard application procedure. Data requests should be submitted to; further details can be found at http://www.nshd.mrc.ac.uk/data.aspx. doi:10·5522/NSHD/Q101; doi:10·5522/NSHD/Q10. Caerphilly Prospective Study data can be accessed following review and approval by the steering committee (https://www.bristol.ac.uk/population-health-sciences/projects/caerphilly/collaboration/). Researchers interested in accessing the Whitehall II data can apply through the Dementias Platform UK (https://www.dementiasplatform.uk) or the Whitehall Scientific Committee (https://www.ucl.ac.uk/psychiatry/research/mental-health-olderpeople/whitehall-ii/data-sharing).

