## Supplementary materials for "Longitudinal associations of psychological distress with subsequent cognitive decline and dementia: a multi-cohort study"

**Figures
Figure S1.** Flow diagram of included studies
**Figure S2.** Pooled association between binary distress and subsequent fluid cognitive levels
**Figure S3.** Pooled association between depressive and anxiety symptoms and subsequent fluid cognitive levels
**Figure S4.** Pooled association between psychological distress and subsequent fluid cognitive change
**Figure S5.** Pooled association between binary psychological distress and subsequent dementia
**Figure S6.** Pooled association between psychological distress and dementia – persistence of symptoms
**Figure S7.** Pooled association between depressive and anxiety symptoms with subsequent dementia

**Tables
Table S1.** Amendments to pre-registered protocol
**Table S2.** Description of datasets
**Table S3.** Summary of datasets and assessments
**Table S4.** Missing covariate data before multiple imputation
**Table S5.** Descriptive statistics for analytic sample versus those excluded from analytic sample

**Supplementary information**

**Figure S1. Flow diagram of included studies**

Participated at baseline:
- ELSA: 11,500
- WHII: 10,308
- CAPS: 2,512
- NSHD: 5,362
- NCDS: 12,535

Had dementia at baseline:
- ELSA: 146
- WHII: 0
- CAPS: N/A
- NSHD: N/A
- NCDS: N/A

No subsequent cognitive information in selected wave for single time point analysis, or in two waves for multilevel models:
- ELSA: 1,907
- WHII: 3,590
- CAPS: 659
- NSHD: 1,111
- NCDS: 6,799

Missing data on mental health at baseline:
- ELSA: 425
- WHII: 119
- CAPS: 673
- NSHD: 2,070
- NCDS: 154

Analytic sample:
- ELSA: 9,022
- WHII: 6,599
- CAPS: 1,180
- NSHD: 2,181
- NCDS: 5,582

With information on mental health at baseline:
- ELSA: 10,929
- WHII: 10,189
- CAPS: 1,839
- NSHD: 3,292
- NCDS: 12,381

No known dementia at baseline:
- ELSA: 11,354
- WHII: 10,308
- CAPS: 2,512
- NSHD: 5,362
- NCDS: 12,535

**Figure S2. Pooled association between binary distress and subsequent fluid cognitive levels**

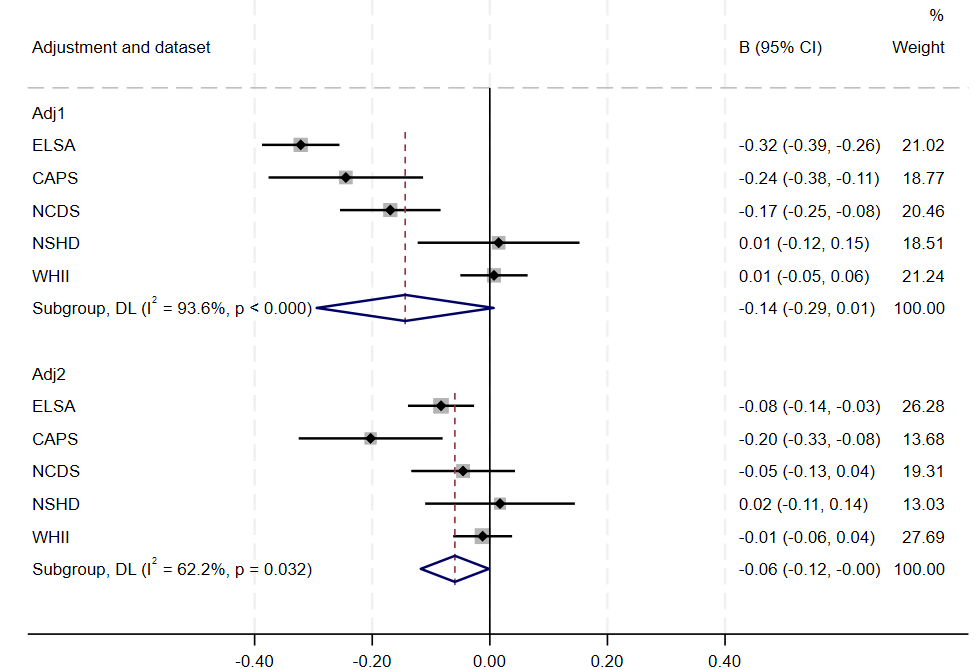

**Figure S3. Pooled association between depressive (left) and anxiety symptoms (right) and subsequent fluid cognitive levels**

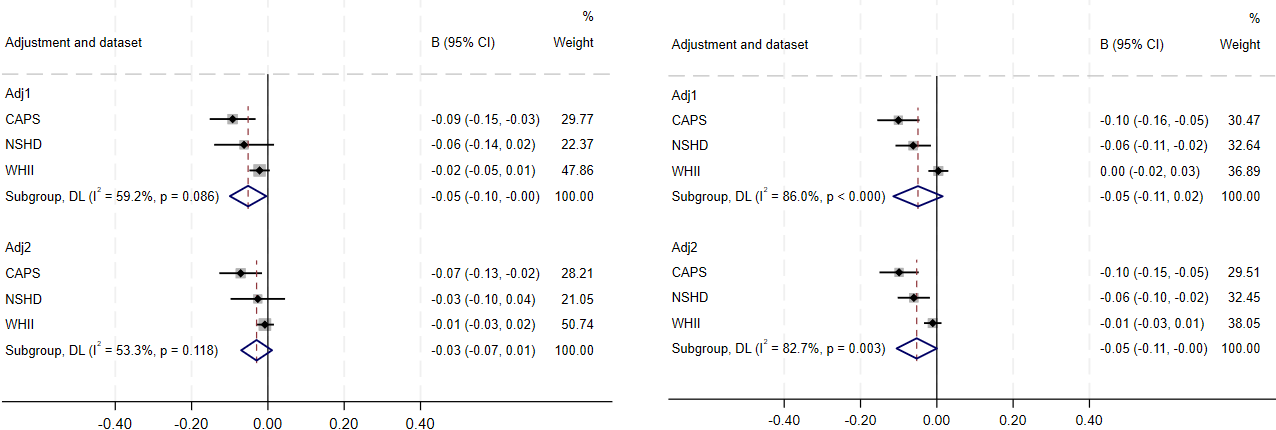

**Figure S4. Pooled association between psychological distress and subsequent fluid cognitive change**

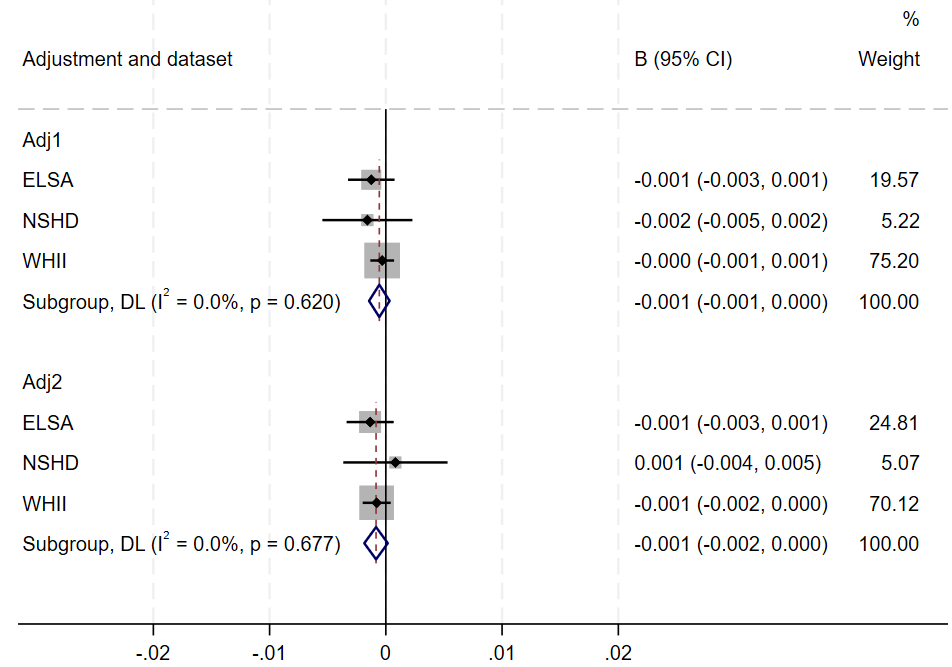

**Figure S5. Pooled association between binary psychological distress and subsequent dementia**

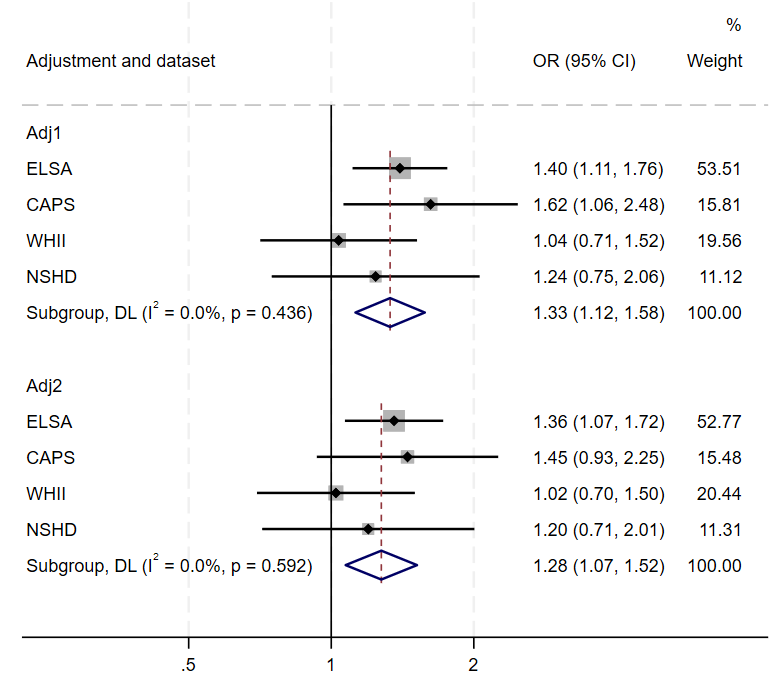

**Figure S6. Pooled association between psychological distress and dementia – persistence of symptoms – partially adjusted (left), fully adjusted (right)**

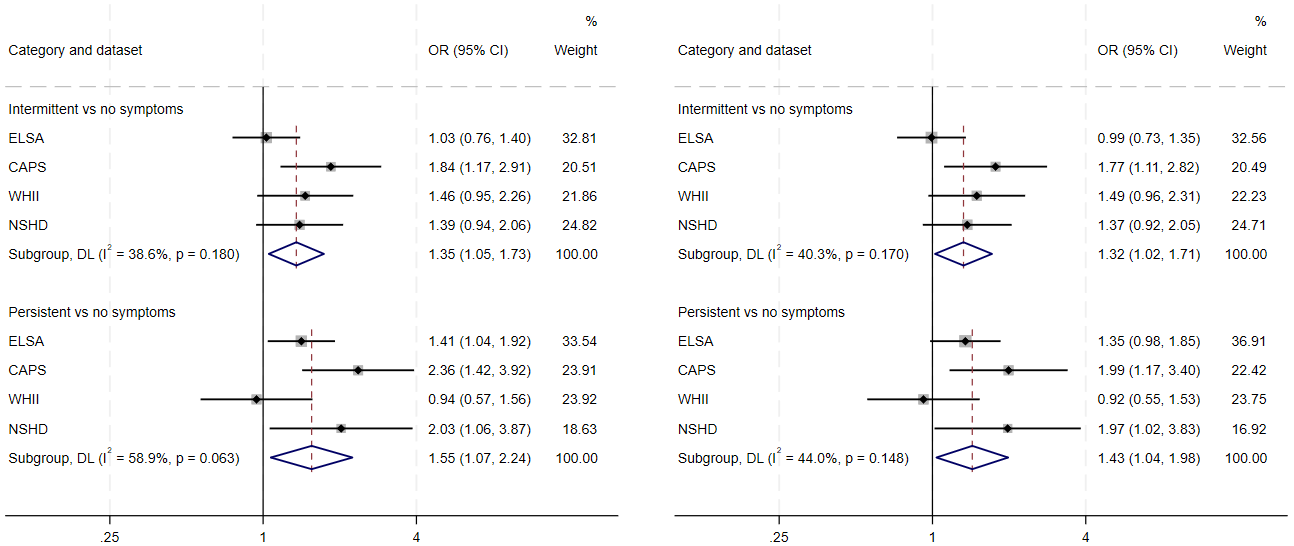

**Figure S7. Pooled association between depressive and anxiety symptoms with subsequent dementia**

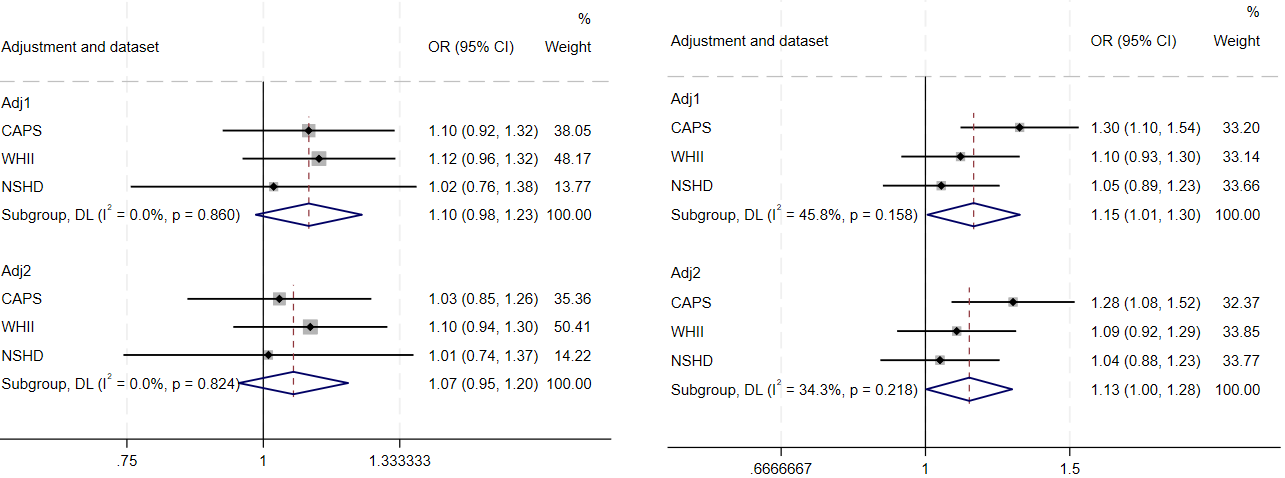

| **Table S1. Amendments to pre-registered protocol** | | |
| --- | --- | --- |
| **Cohort** | **Planned** | **Amendment** |
| Study inclusion | Six longitudinal studies | Five longitudinal studies were analysed due to delays in access to data for one planned cohort. |
| Cognitive outcomes | Associations with both overall composite cognition and individual cognitive domains | Analyses were restricted to overall composite cognition to streamline results and enhance comparability across studies. |
| Statistical modelling approach | Cox proportional hazards regression for dementia outcomes | Logistic regression was used for primary analyses to increase comparability across studies, as several studies assessed dementia at a single time point without information on timing of dementia. The planned analysis on age at psychiatric symptom assessment was retained, but the proposed temporality analyses from Cox models were therefore not undertaken. As a sensitivity analysis, we also examined associations between psychological distress and dementia in ELSA and WHII, where information on timing was available, using Cox regression models. |

| **Table S2. Description of datasets** | |
| --- | --- |
| **Cohort** | **Summary** |
| NSHD | The Medical Research Council (MRC) National Survey of Health and Development (NSHD) is a British birth cohort. The original NSHD sample consisted of 5,362 singleton babies born in one week in March 1946 to married parents in England, Scotland or Wales, stratified by social class. This sample has been followed up twenty-four times since birth. |
| NCDS | The 1958 National Child Development Study (NCDS) follows the lives of 17,415 people born in England, Scotland and Wales in a single week of 1958. It started in 1958 at birth, as the Perinatal Mortality Survey. Surviving members of the cohort were followed up in 1965 (age 7), 1969 (age 11), 1974 (age 16), 1981 (age 23), 1991 (age 33), 1999/2000 (age 41/42), 2004-2005 (age 46-47), 2008-2009 (age 50), 2013 (age 55) and 2020 (age 62). The initial response rate was just over 98% of all births in Great Britain that week, with varying responses in subsequent waves. |
| WHII | The Whitehall II study (WHII) was established to investigate the causes of social inequalities in health. A cohort of 10,308 participants aged 35-55, of whom 3,413 were women and 6,895 were men, was recruited from the British Civil Service in 1985. Since the first wave of data collection, self-completion questionnaires and clinical data have been collected from the cohort every two to five years with a high level of participation. |
| ELSA | The English Longitudinal Study of Ageing (ELSA) is a study of adults aged ≥50 years living in private households in England. In 2002/2003 the original sample was drawn from households that had previously responded to the Health Survey for England (n=11,931). Comparisons of the sociodemographic characteristics of participants against national census results indicate that the sample was broadly representative of the English population. Participants are contacted every two years and data collection consists of a face-to-face interview and self-completion questionnaire. |
| CAPS | The Caerphilly Prospective Study (CAPS) is a population study set up to investigate relationships between lifestyle and other factors with incident cardiovascular disease in men from Caerphilly and surrounding villages in Wales. Phase I began in 1979 until 1983. The cohort was re-examined after approximately 5-years: phase II (1984-1988), phase III (1989-1993), and phase IV (1993-1996). Follow-up research continued from 1997-2016. Phase V (2002-2004) was the 4^th^ follow-up with questionnaires, cognitive and physical measures. The initial sample included 2,512 men. |

| **Table S3. Summary of datasets and assessments** | | | | | | |
| --- | --- | --- | --- | --- | --- | --- |
| **Dataset** | **Baseline n** | **Baseline age** | **Psychological distress (measure, age)** | **Cognition (age/phase/wave)** | **Dementia assessed?** | **Psychiatric symptom severity cut-off** |
| **NSHD** | 5,362 | Birth cohort | PSE (**age 36**); PSF (age 43); GHQ-28 (age 53, 60-64, 69) | Memory, processing speed (age 43, 53, 60-64, 69, 77) | Age 77 | PSE cut-off - ≥4  PSF cut-off - ≥23  GHQ cut-off - ≥5 (binary scoring method) |
| **NCDS** | 17,415 | Birth cohort | Malaise Inventory (**age 23,** 33, 42, 50, 62) | Memory, verbal fluency, processing speed and accuracy (age 50, 62) | No | Cut-off - ≥4 (9-item version) |
| **WHII** | 10,308 | 35-55 | GHQ-30 (**1985-1988**, 1989-1990, 1991-1994, 1997-1999, 2001, 2002-4, 2006, 2007-9, 2012-13, 2015-16, 2019-22) | Verbal fluency, memory, Alice Heim 4-I (1997-99, 2002-04, 2007-09, 2012-13, 2015-16, 2019-22) | All waves | Cut-off - ≥5 (binary scoring method) |
| **ELSA** | 11,931 | 50-99+ | CES-D (**2002-2003**; 2004-2005; 2006-2007; 2008-2009; 2010-2011; 2012-2013; 2014-2015; 2016-2017; 2018-2019; 2021-2023) | Memory, verbal fluency, orientation (all waves; except verbal fluency unavailable in wave 6) | All waves | Cut-off - ≥4 |
| **CAPS** | 2,512 | 45-59 | GHQ-30 (**1979-1983**; 1984-1988; 1989-1993; 1993-1997) | CAMCOG (1989-1993; 1993-1997; 2002-2004) | Phase 5 | Cut-off - ≥5 (binary scoring method) |

| **Table S4. Missing covariate data in analytic sample before multiple imputation** | | | | | |
| --- | --- | --- | --- | --- | --- |
| **Covariates** | **ELSA** | **CAPS** | **WHII** | **NSHD** | **NCDS** |
| **Overall N** | 9,022 | 1,180 | 6,599 | 2,181 | 5,582 |
| **Baseline age, mean (standard deviation (SD))** | 64.1 (9.7) | 51.5 (4.4) | 44.3 (5.9) | 36 | 23 |
| *Missing* | - | - | - | - | - |
| **Sex** |  |  |  |  |  |
| Male | 4,034 (44.7%) | 1,180 (100%) | 4,712 (71.4%) | 1,041 (47.7%) | 2,691 (48.2%) |
| Female | 4,988 (55.3%) | - | 1,887 (28.6%) | 1,140 (52.3%) | 2,891 (51.8%) |
| *Missing* | - | - | - | - | - |
| **Marital status** |  |  |  |  |  |
| Not married or cohabiting | 1,924 (21.3%) | 89 (7.5%) | 1,550 (23.5%) | 217 (9.9%) | 3,075 (55.1%) |
| Married (/cohabiting) | 7,098 (78.7%) | 1,091 (92.5%) | 5,035 (76.3%) | 1,964 (90.1%) | 2,507 (44.9%) |
| *Missing* | - | - | 14 (0.2%) | - | - |
| **Education** |  |  |  |  |  |
| Less than o-level or equivalent | 3,978 (44.1%) | 718 (60.9%) | 1.383 (21.0%) | 830 (38.1%) | 1,004 (17.8%) |
| O-level or equivalent | 4,269 (47.3%) | 425 (36.0%) | 3,534 (53.6%) | 1,260 (57.8%) | 3,774 (67.6%) |
| International/other | 772 (8.6%) | - | - | - | - |
| *Missing* | <10 (<0.5%) | 37 (3.1%) | 1,682 (25.5%) | 91 (4.2%) | 804 (14.4%) |
| **Occupational social class** |  |  |  |  |  |
| Manual | 3,566 (39.5%) | 705 (59.8%) | Administrative: 2,251 (34.11%) | 918 (42.1%) | 2,034 (36.4%) |
| Non-manual | 5,105 (58.6%) | 464 (39.3%) | Prof/exec: 3,349 (50.8%) | 1,159 (53.1%) | 3,321 (59.5%) |
| Other | - | - | Clerical/support: 999 (15.1%) | 56 (2.6%) | - |
| *Missing* | 351 (3.9%) | 11 (0.9%) | - | 48 (2.2%) | 227 (4.1%) |
| **Chronic health conditions** |  |  |  |  |  |
| None | 6,704 (74.3%) | 1,021 (86.5%) | 5,904 (89.5%) | 2,100 (96.3%) | 5,398 (96.7%) |
| One or more | 2,297 (25.5%) | 159 (13.5%) | 695 (10.5%) | 78 (3.6%) | 182 (3.3%) |
| *Missing* | 21 (0.2%) | - | - | <10 (<0.5%) | <10 (<0.5%) |
| **Smoking status** |  |  |  |  |  |
| Non-smoker | 3,279 (36.3%) | 238 (20.2%) | 3,452 (52.3%) | 698 (32.0%) | 1,821 (32.6%) |
| Previous smoker | 4,162 (46.1%) | 364 (30.9%) | 2,221 (33.7%) | 860 (39.4%) | 1,761 (31.5%) |
| Current smoker/smoked regularly | 1,581 (17.5%) | 573 (48.6%) | 923 (14%) | 621 (28.5%) | 2,000 (35.8%) |
| *Missing* | - | <10 (<0.5%) | <10 (<0.5%) | <10 (<0.5%) | - |
| **Alcohol consumption** |  |  |  |  |  |
| Low | 2,635 (29.2%) | 223 (18.9%) | 970 (14.7%) | - | 963 (17.3%) |
| Moderate | 3,768 (41.8%) | 708 (60.0%) | 4,417 (66.9%) | - | 3,446 (61.7%) |
| High | 2,618 (29.0%) | 222 (18.8%) | 1,164 (17.6%) | - | 1,170 (20.1%) |
| *Missing* | <10 (<0.5%) | 27 (2.3%) | 48 (0.7%) |  | <10 (<1%) |
| **Average units alcohol per day – mean (SD)** | - | - | - | 1.8 (2.5) | - |
| *Missing* | - | - | - | 452 (20.7%) | - |
| **Physical activity** |  |  |  |  |  |
| Low | 2,635 (29.2%) | 608 (51.5%) | 2,173 (32.9%) | 738 (33.8%) | 3,091 (55.4%) |
| Moderate | 3,769 (41.8%) | 218 (18.5%) | 2,698 (40.9%) | 597 (27.4%) | 1,657 (29.7%) |
| High | 2,618 (29.0%) | 280 (23.7%) | 1,631 (24.7%) | 785 (36.0%) | 829 (14.9%) |
| *Missing* | <10 (<0.5%) | 74 (6.3%) | 96 (1.5%) | 61 (2.8%) | <10 (<1%) |

| **Table S5. Descriptive statistics for analytic sample versus those excluded from analytic sample** | | | | | | | | | | |
| --- | --- | --- | --- | --- | --- | --- | --- | --- | --- | --- |
| **Covariates** | **ELSA (analytic sample) n=9,022** | **ELSA (N varies – total excluded n=3,077)** | **CAPS (analytic sample) n=1,180** | **CAPS (N varies – total excluded n=1,332)** | **WHII (analytic sample) n=6,599** | **WHII (N varies – total excluded n=3,709)** | **NSHD (analytic sample) 2,181** | **NSHD (N varies – total excluded n=3,181)** | **NCDS (analytic sample) N=5,582** | **NCDS (N varies – total excluded n=6,952)** |
| **Baseline age, mean (standard deviation (SD))** | 64.1 (9.7) | 64.1 (13.8) | 51.5 (4.4) | 52.7 (4.4) | 44.3 (5.9) | 46.1 (6.2) | 36 | 36 | 23 | 23 |
| **Sex** |  |  |  |  |  |  |  |  |  |  |
| Male | 4,034 (44.7%) | 1,301 (42.3%) | 1,180 (100%) | 1,332 (100%) | 4,712 (71.4%) | 2,183 (58.9%) | 1,041 (47.7%) | 1,773 (55.8%) | 2,691 (48.2%) | 3,574 (50.4%) |
| Female | 4,988 (55.3%) | 1,776 (57.5%) | - | - | 1,887 (28.6%) | 1,526 (41.1%) | 1,140 (52.3%) | 1,406 (44.2%) | 2,891 (51.8%) | 3,378 (48.6%) |
| **Marital status** |  |  |  |  |  |  |  |  |  |  |
| Not married (or cohabiting) | 1,924 (21.3%) | 632 (20.5%) | 89 (7.5%) | 206 (15.5%) | 1,553 (23.5%) | 1,112 (30.0%) | 217 (9.9%) | 200 (17.5%) | 3,075 (55.1%) | 3,868 (55.7%) |
| Married (or cohabiting) | 7,098 (78.7%) | 2,445 (79.5%) | 1,091 (92.5%) | 1,126 (84.5%) | 5,046 (76.5%) | 2,573 (69.4%) | 1,964 (90.1%) | 940 (82.5%) | 2,507 (44.9%) | 3,080 (44.3%) |
| **Education** |  |  |  |  |  |  |  |  |  |  |
| Less than o-level or equivalent | 3,981 (44.1%) | 1,612 (52.9%) | 741 (62.8%) | 305 (75.5%) | 1,805 (27.4%) | 1,111 (40.7%) | 873 (40%) | 1,287 (55.0%) | 1,205 (21.6%) | 1,430 (32.7%) |
| O-level or equivalent | 4,269 (47.3%) | 1,190 (39.1%0 | 439 (37.2%) | 99 (24.5%) | 4,794 (72.6%) | 3,534 (71.9%) | 1,308 (60%) | 1,053 (45.0%) | 4,377 (78.4%) | 2,946 (67.3%) |
| International/other | 772 (8.6%) | 243 (8.0%) | - | - | - |  | - | - | - | - |
| **Occupational social class** |  |  |  |  |  |  |  |  |  |  |
| Manual | 3,732 (41.4%) | 1,450 (50.9%) | 715 (60.6%) | 963 (75.5%) | Clerical/support: 999 (15.1%) | 1,338 (36.1%) | 944 (43.3%) | 1,009 (42.0%) | 2,135 (38.2%) | 3,290 (49.5%) |
| Non-manual | 5,290 (58.6%) | 1,400 (49.1%) | 465 (39.4%) | 313 (24.5%) | Administrative: 2,251 (34.11%) | 777 (21.0%) | 1,181 (54.1%) | 795 (33.1%) | 3,447 (61.8%) | 3,361 (50.5%) |
| Other | - | - | - | - | Professional/executive: 3,349 (50.8%) | 1,594 (43.0%) | 56 (2.6%) | 697 (24.9%) | - | - |
| **Chronic health conditions** |  |  |  |  |  |  |  |  |  |  |
| None | 6,725 (74.5%) | 2,141 (69.9%) | 1,021 (86.5%) | 1,022 (76.7%) | 5,904 (89.5%) | 3,162 (85.3%) | 2,103 (96.4%) | 1,056 (92.8%) | 5,400 (96.7%) | 6,552 (94.3%) |
| One or more | 2,297 (25.5%) | 922 (30.1%) | 159 (13.5%) | 310 (23.3%) | 695 (10.5%) | 547 (14.8%) | 78 (3.6%) | 82 (7.2%) | 182 (3.3%) | 394 (5.7%) |
| **Smoking status** |  |  |  |  |  |  |  |  |  |  |
| Non-smoker | 3,279 (36.3%) | 1,007 (34.9%) | 239 (20.3%) | 154 (11.6%) | 3,453 (52.3%) | 1,679 (45.4%) | 699 (32.0%) | 288 (25.3%) | 1,821 (32.6%) | 2,041 (29.4%) |
| Previous smoker | 4,162 (46.1%) | 1,299 (45.0%) | 364 (30.8%) | 359 (27.1%) | 2,222 (33.7%) | 1,060 (28.6%) | 861 (39.5%) | 344 (30.2%) | 1,761 (31.5%) | 1,802 (26.0%) |
| Current smoker/smoked regularly | 1,581 (17.5%) | 580 (20.1%) | 577 (48.9%) | 814 (61.3%) | 924 (14%) | 963 (26.0%) | 621 (28.5%) | 506 (44.5%) | 2,000 (35.8%) | 3,101 (44.7%) |
| **Alcohol consumption** |  |  |  |  |  |  |  |  |  |  |
| Low | 2,635 (29.2%) | 1,075 (37.3%) | 228 (19.3%) | 278 (22.3%) | 973 (14.7%) | 903 (24.7%) | - | - | 963 (17.3%) | 1,433 (20.6%) |
| Moderate | 3,769 (41.8%) | 1,104 (38.3%) | 725 (61.4%) | 721 (57.8%) | 4,456 (67.5%) | 2,189 (59.8%) | - | - | 3,447 (61.8%) | 4,116 (59.3%) |
| High | 2,618 (29.0%) | 704 (24.4%) | 227 (19.2%) | 248 (19.9%) | 1,170 (17.7%) | 571 (15.6%) | - | - | 1,172 (21.0%) | 1,394 (20.1%) |
| **Average units alcohol per day – mean (SD)** | - | - | - | - | - | - | 1.8 (2.5) | 2.1 (3.1) | - | - |
| **Physical activity** |  |  |  |  |  |  |  |  |  |  |
| Low | 2,635 (29.2%) | 996 (34.5%) | 650 (55.1%) | 734 (59.8%) | 2,213 (33.5%) | 1,453 (40.6%) | 756 (34.7%) | 481 (43.4%) | 3,095 (55.4%) | 4,272 (61.6%) |
| Moderate | 3,769 (41.8%) | 1,248 (43.3%) | 232 (19.7%) | 188 (15.3%) | 2,741 (41.5%) | 1,484 (41.5%) | 614 (28.2%) | 294 (26.5%) | 1,658 (29.7%) | 1,707 (24.6%) |
| High | 2,618 (29.0%) | 640 (22.2%) | 298 (25.3%) | 305 (24.9%) | 1,645 (24.9%) | 643 (18.0%) | 811 (37.2%) | 334 (30.1%) | 829 (14.9%) | 960 (13.8%) |

| **Supplementary Information – Psychological distress** | | |
| --- | --- | --- |
| **Study** | **Measures** | **Age/year/wave (for present study)** |
| **Psychological distress** | | |
| NSHD | - Present State Examination (PSE). Standardised interview administered by trained nurses – assesses low mood, anxiety and phobia symptoms compared to one month before interview. - Psychiatric Symptom Frequency scale. 19-item scale measuring current and recent (in the last 12 months) anxiety and depression. - GHQ-28 – self-administered screening questionnaire which assesses recent symptoms (past few weeks) of common mental disorder. Each item is scored using a 4-point Likert scale and recoded into binary values. | Age 36  Age 43  Ages 53, 60-64, 69 |
| NCDS | Malaise Inventory Scale. Self-completion items combined to assess emotional disturbance and associated somatic symptoms. For comparison across waves, we used the 9-item version available across all waves. Each item is scored from 0-1 and summed to generate an overall score. | Age 23, 33, 42 & 50 |
| Whitehall II | General Health Questionnaire-30 (GHQ-30), an established questionnaire assessing psychological distress, with response items to 30 questions ranging from not at all to much more than usual. | 1985, 1989, 1991, 1997, 2001, 2003, 2006, 2007, and 2012 |
| ELSA | 8-item Center for Epidemiologic Studies Depression Scale. Yes/no for each item. Items coded so that higher scores indicate more depressive symptoms. | Waves 1-10 |
| CAPS | General Health Questionnaire-30 (GHQ-30). | Phases 2-4 |
| **Depressive symptoms (where separate subscales available)** | | |
| NSHD | GHQ depression items - 4 items: Been thinking of yourself as a worthless person; Felt that life is entirely hopeless; Felt that life isn’t worth living; Found you couldn’t do anything because your nerves were too bad. | Age 53, 60-64, 69 |
| NCDS | N/A | Age 23, 33, 42 & 50 |
| Whitehall II | GHQ depression items – as above. | 1985, 1989, 1991, 1997, 2001, 2003, 2006, 2007, and 2012 |
| ELSA | N/A | Waves 1-10 |
| CAPS | GHQ depression items – as above. | Phases 2-4 |
| **Anxiety symptoms (where separate subscales available)** | | |
| NSHD | GHQ anxiety subscale – 5 items - Lost much sleep over worry; Felt constantly under strain; Been getting scared or panicky for no good reason; Found everything getting on top of you; Been feeling nervous and strung up all the time. | Ages 53, 60-64 and 69 |
| NCDS | N/A | N/A |
| Whitehall II | GHQ anxiety subscale – as above. | 1985, 1989, 1991, 1997, 2001, 2003, 2006, 2007, 2012, 2015 |
| ELSA | N/A | N/A |
| CAPS | GHQ anxiety subscale – as above. | Phases 2-4 |

| **Supplementary Information - overlapping self-report measures within NSHD – from McElroy et al. 2020** | | |
| --- | --- | --- |
| **Symptom** | **PSF (NSHD – age 43)** | **GHQ-28 item (NSHD – age 53, 60-64, 69)** |
| **Low mood** | 2. Have you been in low spirits or felt miserable | 17. Been able to enjoy your normal day-to-day activities |
| **Fatigue** | 14. Have there been days when you tired out very easily? | 2. Been feeling in need of a good tonic |
| **Tense/stressed** | 14. Have there been days when you tired out very easily? | 23. Been feeling nervous and strung-up all the time |
| **Sleep problems** | 9. Have you had trouble getting off to sleep | 8. Lost much sleep over worry |
| **Panic** | 8. Have you been in situations when you felt shaky or sweaty or your heart pounded or you could not get your breath | 19. Been getting scared or panicky for no good reason |
| **Hopelessness** | 16. Have you had the feeling that the future does not hold much for you? | 22. Felt that life is entirely hopeless |
| **Health anxiety** | 11. Have you been frightened or worried about becoming ill or about dying? | 4. Felt that you are ill |

| **Supplementary Information – variable recoding from PSF and GHQ (Mcelroy et al., 2020)** | | | |
| --- | --- | --- | --- |
| **PSF** |  | **GHQ** |  |
| **Value** | **Label** | **Value** | **Label** |
| 0    1 | Never    Occasionally | 1    2 | Not at all    No more than usual |
| 2 | Sometimes | 3 | Rather more than usual |
| 3 | Quite often | 4 | Much more than usual |
| 4 | Very often |  |  |
| 5 | Always |  |  |

Note: Line indicates placement of binary split. Above the line coded as 0 (no symptoms), below the line coded as 1 (symptom present)

| **Supplementary Information - cognitive outcomes** | |
| --- | --- |
| **Dataset** | **Description of variable** |
| NSHD | Cognition was assessed in 1999, 2006-2010, 2014-2015, and 2023, when participants were aged 53, 60-64, 68-69 and 73 years. To assess immediate recall, participants were shown 15 words and asked to write down as many as possible from memory in any order, with different word lists used to minimise practice effects. Tests were completed three times, and scores from the first test were included in the present study. Processing speed was tested using a letter cancellation test where participants were given a page of random letters from the alphabet set out in 26 rows and 30 columns and asked to cross out as many ‘Ps’ and ‘Ws’ as possible within one minute (65 target letters in total). |
| NCDS | We focused on cognitive outcomes assessed at age 62 years. We computed a composite cognitive score based on domains of memory, assessed with a word-recall test of immediate and delayed recall; a verbal fluency task in which respondents were asked to name as many animals as possible within one minute; and a letter cancellation task to assess executive functioning, with participants asked to cross as many of the letters ‘P’ and ‘W’ as they could spot in the list of letters within one minute. |
| Whitehall II | Participants completed cognitive testing in 1997-99, 2002-04, 2007-09, 2012-13, 2015-16, and 2019-2022. We generated composite cognitive scores from a verbal fluency task, where participants were asked to write down as many words as possible beginning with ‘S’ and as many animals as possible in 1 minute; a measure of short-term verbal memory where participants were presented with 20 1- or 2-syllable words at 2-second intervals, and had 2 minutes to recall as many words as possible in writing; and the Alice Heim 4-I test which assesses mathematical and verbal reasoning |
| ELSA | In ELSA, assessments of verbal fluency, recall and orientation took place between waves 1-10, although verbal fluency was not available in wave 6. Composite cognition was computed from a verbal fluency task in which individuals were required to verbally name as many animals as possible in one minute, and memory tests in which participants had to recall a list of 10 unrelated words both immediately and after a delay. |
| CAPS | Composite cognition was indexed between phases 3-5 using the Cambridge Cognition Examination (CAMCOG), a neuropsychological test battery covering a range of cognitive skills and functions, including orientation, language, memory, praxis, attention, abstract thinking, perception and calculation. In phases 3 and 4, an adapted version was used excluding the three easiest items due to the younger population, reducing the number of items to 57 and the maximum score to 104. In Phase 5, scores ranged from 0 to 102. |

| **Supplementary Information - dementia outcome** | |
| --- | --- |
| **Dataset** | **Description of variable** |
| Whitehall II | All-cause dementia was obtained from the national Hospital Statistics database using International Classification of Diseases 10^th^ Edition (ICD-10) codes ICD-10 (F00-F04, G30, G31) via HES, with 78% sensitivity & 92% specificity. |
| ELSA | Self-reported physician-diagnosis of dementia or Alzheimer’s disease. Additional cases ascertained using Informant Questionnaire on Cognitive Decline in the Elderly (IQCODE), consisting of 16 items asking informant to report the ability of an individual compared with 10 years ago to perform various function, such as remembering names of family and friends, with five-point scores for each item ranging from much improved to much worse. Cut-off of 3.5 to define dementia. High specificity and sensitivity. |
| CAPS | At Phase V (men aged 68–82 years), poor performance on the Cambridge Cognition Exam (CAMCOG) was used to select men for detailed clinical assessment. Subjects were classified as having normal cognition, cognitive impairment no dementia (CIND), or dementia. |
| NSHD | The AD8 was administered in 2023 (age 77 years). The AD8 is a short interview designed to be administered to participants’ informants to distinguish those with very mild dementia from those without dementia. A validity study showed strong correlation between AD8 scores, Clinical Dementia Rating domains and performance on neuropsychological tests (Galvin et al., 2006). The interview includes 8-items in which informants are asked wither there have been changes over the last several years caused by cognitive problems in the following areas: problems with judgment; less interest in hobbies/activities; repeats the same thing over and over; trouble learning how to use a tool, appliance or gadget; forgets correct month or year; trouble handling complicated financial affairs; trouble remembering appointments; daily memory problems. Each item is coded as no change or yes change, and a total score is derived, with a cut-off score of two commonly used to screen for cognitive impairment and early dementia symptoms. |
| NCDS | / |

| **Supplementary Information - covariates** | | |
| --- | --- | --- |
| **Dataset** | **Description of variable** | **Response** |
| **Marital status** (married and/or living with a partner; unmarried/unmarried and living alone) | | |
| NSHD | What is your current marital status? | Single and not living with a partner; single and living with a partner; married; widowed; separated and not living with a partner; divorced and not living with a partner; separated or divorced and living with a partner |
| NCDS | Present marital status | Single; legally married; separated; divorced; widowed |
| Whitehall II | Marital status | Married; cohabit; single; divorced; widowed |
| ELSA | Current legal marital status | Single, married, first and only marriage; remarried, second or later marriage; legally separated; divorced; widowed  Household size (numeric) |
| CAPS | Marital status | Married; single; widowed; divorced; separated |
| **Education** (GCSE/O-level or equivalent; less than GCSE/O-level) | | |
| NSHD | Highest educational attainment up to 26 years | None; vocational course, proficiency only; Sub GCE or Burnham C; GCSE O-Level or Burnham C; GCSE A-Level or Burnham B; Burnham A2; 1st Degree or graduate equivalent; Higher degree, masters; Higher degree, doctorate |
| NCDS | Highest qualification gained at age 23 | No qualification; CSE 2-5/equivalent NVQ1; O Level equivalent NVQ2; A level/equivalent NVQ3; Higher qualification NVQ4; Degree/higher NVQ5,6 |
| Whitehall II | Type of school/college last attended Education level | Secondary; university/polytechnic; nursing; college; other Up to 16; 17-18; over 18 |
| ELSA | Highest educational attainment | NVQ4/NVQ5/Degree or equiv; Higher ed below degree; NVQ3/GCE A Level equiv; NVQ2/GCE O Level equiv; NVQ1/CSE other grade equiv; Foreign/other; No qualification |
| CAPS |  | School certificate or CSE grade 1 or GCE O level or Scottish(SCE) lower or City & Guilds Craft (Ordinary level); GCE A level or Higher certificate or Matriculation or Scottish(SCE) higher; overseas school leaving exam or certificate; ONC or OND or City & Guilds Advanced or Final level; HNC or JNC or City & Guilds Full Technological Certificate; RSA or other clerical and commercial; teacher training; nursing qualification; professional qualification; degree including higher degree; other qualification |
| **Occupational class** (manual; non-manual) | | |
| NSHD | Social class of head of household | Armed forces; I professional; II intermediate; III skilled (non-manual); IIIM skilled manual; IV partly skilled; V unskilled; never worked full time; unmarried woman (own social class) |
| NCDS | Adult socio-economic position | Professional/intermediate; other non-manual; skilled manual; other manual |
| Whitehall II | Occupational social class | Professional, managerial, skilled nonmanual, skilled manual, partly skilled, nonskilled |
| ELSA | Occupational class  Three-class National Statistics – Socioeconomic Classification Scheme | I – Professional; II – Managerial technical; IIIN – skilled non-manual; IIIM – skilled manual; IV – semi-skilled manual; V – unskilled manual; armed forces |
| CAPS | What is your present work? | General's classification of I, II, III non-manual, III manual, IV and V. |
| **Long-term health conditions** (none; one or more) | | |
| NSHD | Self-reported diabetes, heart trouble or high blood pressure | Yes/no option for each diagnosis |
| NCDS | Longstanding illness or disability | Yes; no |
| Whitehall II | Number of chronic conditions, including diabetes, coronary heart disease and stroke – based on self-reported physician diagnosis, use of diabetes medication or HES record | Yes/no option for each diagnosis |
| ELSA | Self-report doctor diagnosed: angina, heart attack/myocardial infarction, congestive heart failure, heart murmur, abnormal heart rhythm, diabetes or high blood sugar, stroke/cerebral vascular disease. | Yes/no option for each diagnosis |
| CAPS | Have you ever had any of the following illnesses? Stroke; heart attack or coronary thrombosis; diabetes  LSHTM Rose Angina | Yes/no option for each diagnosis  No; Angina grade I; Angina grade II |
| **Physical activity** (low; moderate; high) | | |
| NSHD | At age 36, participation in leisure time physical activity was ascertained using a modified validated Minnesota leisure time physical activity questionnaire assessing how often people took part in a range of physical activities per month. Frequency of participation in physical activity in the past 4 weeks, including: football, gym, golf, jogging, badminton, bowls, climbing; yoga; cricket; rowing; running; sailing; squash, swimming; table tennis; tennis; home keep fit exercises; skiing; volleyball; dancing; riding; basketball; weight training; fishing; ballroom dancing; scuba diving; moving to music | Continuous  *Grouped into inactive (no participation in leisure time physical activity per month); moderately active (one to five times per month), or active (six or more times per month).* |
| NCDS | At age 23, participants were asked about activities during leisure time over the past 4 weeks. Participants were asked whether they had played sport of any kind, including keep fit, yoga and similar exercise. | 5 times a week or more often/3 or 4 times a week; once or twice a week/2 or 3 times in the last 4 weeks; once in the last 4 weeks/not at all in the last 4 weeks |
| Whitehall II | Frequency of participation in vigorous and moderate exercise per week | Often/3+ times a week; sometimes/1-2 times a week; seldom/1-3 times a month; never/hardly ever |
| ELSA | Do you take part in sports or activities that are moderately energetic? Do you take part in sports or activities that are vigorous? | More than once a week; once a week; one to three times a month; hardly ever or never |
| CAPS | How many minutes of heavy work or exercise in leisure time in previous 7 days | Low=0-10 minutes; moderate=10-120 minutes; high=121 minutes and above |
| **Smoking** (current smoker; ex-smoker; or never smoked) | | |
| NSHD | Smoking status | Current smoker; ex-smoker; never smoked |
| NCDS | Ever smoked cigarettes, cigar or pipe Current cigarette smoking | Yes; no |
| Whitehall II | Currently smoke cigarettes Ever smoked cigarettes | Yes; no Yes; no |
| ELSA | Smoking status | Never smoked; ex-smoker – occasional; ex-smoker – regular; ex-smoker but don't know frequency; current smoker |
| CAPS | Do you smoke at all? Have you ever been a regular smoker? | No; yes cigarettes; yes cigars; yes pipe; yes present smoker |
| **Alcohol intake** (low; moderate; or high) | | |
| NSHD | Average units of alcohol per day | Continuous |
| NCDS | Frequency of drinking | Most days; 1-2 times a week; less often; special occasion; never drink |
| Whitehall II | Measures of spirits, glasses of wine and pints of beer in past week Whether or not consumed alcohol in past week | Continuous Yes; no Grouped into none, low to moderate, or high by sex based on UK government guidelines |
| ELSA | In the past 12 months have you taken an alcoholic drink? | Twice a day or more/daily or almost daily; once or twice a week/once or twice a month; special occasions only/not at all |
| CAPS | Do you take some drinks containing alcohol? | Most days/every day; weekends and occasionally in the week/weekends only/not every week; never/special occasions only |
| **Baseline or childhood cognition** | | |
| NSHD | At 8 years, participants took tests of verbal and non-verbal ability devised by the National Foundation for Educational Research, and administered by teachers or other trained personnel. These tests were as follows: (1) reading comprehension (selecting appropriate words to complete 35 sentences), (2) word reading (ability to pronounce 50 words), (3) vocabulary (ability to explain the meaning of these 50 words) and (4) picture intelligence, consisting of a 60-item non-verbal reasoning test. Scores for each test were standardised to the whole sample, then summed to create a total score representing overall cognitive ability at this age. | Continuous score |
| NCDS | Childhood cognition was assessed at age 11, using a general ability test administered at the child’s school. | Continuous score |
| Whitehall II | - | - |
| ELSA | As described above in cognition section | Continuous score |
| CAPS | - | - |
